# Longitudinal Changes of Cardiac and Aortic Imaging Phenotypes Following COVID-19 in the UK Biobank Cohort

**DOI:** 10.1101/2021.11.04.21265918

**Authors:** Wenjia Bai, Betty Raman, Steffen E. Petersen, Stefan Neubauer, Zahra Raisi-Estabragh, Nay Aung, Nicholas C Harvey, Naomi Allen, Rory Collins, Paul M. Matthews

## Abstract

Case studies conducted after recovery from acute infection with SARS-CoV-2 have frequently identified abnormalities on CMR imaging, suggesting the possibility that SARS-CoV-2 infection commonly leads to cardiac pathology. However, these observations have not been able to distinguish between associations that reflect pre-existing cardiac abnormalities (that might confer a greater likelihood of more severe infection) from those that arise as consequences of infection. To address this question, UK Biobank volunteers (n=1285; 54.5% women; mean age at baseline, 59.8 years old; 96.3% white) who attended an imaging assessment including cardiac magnetic resonance (CMR) before the start of the COVID-19 pandemic were invited to attend a second imaging assessment in 2021. Cases with evidence of previous SARS-CoV-2 infection were identified through linkage to PCR-testing or other medical records, or a positive antibody lateral flow test; n=640 in data available on 22 Sep 2021) and were matched to controls with no evidence of previous infection (n=645). The majority of these infections were milder and did not involve hospitalisation. Measures of cardiac and aortic structure and function were derived from the CMR images obtained on the cases before and after SARS-CoV-2 infection from images for the controls obtained over the same time interval using a previously validated, automated algorithm. Cases and controls had similar cardiac and aortic imaging phenotypes at their first imaging assessment. Changes between CMR imaging measures in cases before and after infection were not significantly different from those in the matched control group. Additional adjustment for comorbidities made no material difference to the results. While these results are preliminary and limited to imaging metrics derived from automated analyses, they do not suggest clinically significant persistent cardiac pathology in the UK Biobank population after generally milder (non-hospitalised) SARS-CoV-2 infection.

## Introduction

Pulmonary disease arising from SARS-CoV-2 (severe acute respiratory syndrome coronavirus-2), which was first recognised in Wuhan, China in December 2019, now has been well described^1^. Like other coronaviruses, pulmonary symptoms and signs dominate its clinical expression as COVID-19 (2019 Corona Virus Disease). However, many clinical symptoms and signs are thought to be related to extrapulmonary organ involvement. Because SARS-CoV-2 enters cells via angiotensin converting enzyme 2, which is expressed widely in vascular endothelia and in some other organs, extrapulmonary manifestations could be a consequence either of direct viral effects on extrapulmonary tissues or consequences of the sometimes severe systemic immunopathological and thrombotic sequalae^2^.

COVID-19 has been associated with multiple symptoms referable to the heart including shortness of breath, new arrythmias, chest pain and non-specific symptoms, as well as non-specific symptoms of fatigue and post-exertional malaise that persist after resolution of the acute disease (“Long COVID”)^3^. These have raised concerns for cardiac injury with infection. There is little definitive evidence for this, although there have been multiple case reports, as summarised in a recent review of CMR findings for patients with COVID-19^4^. In brief, an early report from Wuhan, China described imaging evidence of cardiac abnormalities in 58% of retrospectively recruited patients who recovered from COVID-19^5^. As many as 78% of a younger German cohort imaged after SARS-CoV-2 infection showed abnormal CMR findings, such as increased cardiac muscle MRI T1 that had a modest relationship to elevated high-sensitivity troponin T^6^. Approximately one-quarter of a group of 58 hospitalised COVID-19 patients with persistent limitations to exercise tolerance who were imaged 2-3 months after discharge from the hospital had regional cardiac MRI T1 relaxation times that were increased relative to matched controls, a pattern associated with myocarditis^7^. Another study of a small group of athletes after recovery from COVID showed that 46% had late gadolinium enhancement on cardiac MRI, with 15% showing cardiac regional T1 and T2 relaxation times that were increased related to healthy control values^8^. However, all of these reports been based on studies of patients *after* their hospitalisations with severe COVID-19. None of the studies included images obtained on the same subjects *before and after* infection with SARS-CoV-2. All of these studies therefore potentially are affected by “reverse causation bias”, whereby increased susceptibility to infection because of pre-existing disease cannot be distinguished from cardiac abnormalities caused by SARS-CoV-2 infection.

Other evidence suggests that persistent cardiac injury after SARS-CoV-2 infection is unlikely. In an autopsy series of 40 COVID-19 cases, while non-specific pathological signs of acute and chronic injury were found, evidence for myocarditis was identified in only one^9^. CMR imaging changes also have low biological specificity in the context of people hospitalised with severe diseases^10,11^. The question of whether potential pathology identified by cardiac imaging is independently clinically significant after SARS-CoV-2 infection demands prospective, longitudinal observations of people imaged before and after COVID-19 across the full disease spectrum.

UK Biobank is a research resource based on prospective, longitudinal follow-up of 500,000 UK volunteers that recently has performed the first longitudinal CMR study of a cohort of people imaged both before and after SARS-CoV-2 infection^12^. This work is based on the UK Biobank Imaging Enhancement Study, which was established to comprehensively image a subset of up to 100,000 participants with magnetic resonance imaging (MRI) of the brain, heart and abdomen, dual-energy x-ray absorptiometry (DEXA) and carotid ultrasound^13^. Between 2015 and March 2020, 50,000 of the UK Biobank Imaging Study participants attended one of its three dedicated imaging assessment centres in Cheadle, Newcastle and Reading for a range of imaging phenotyping that included 1.5 T MRI of the heart (measures of ventricular function such as stroke volume, ejection fraction and myocardial strain and thoracic aortic size and distensibility). After the onset of the COVID pandemic, a nested case-control study was developed with the aim of re-imaging (at the same imaging centre and using an identical imaging protocol) about 100-cases with evidence of previous SARS-CoV-2 infection matched to 1,000 controls without evidence of previous infection, both of whom had undergone imaging assessment prior to the SARS-CoV-2 pandemic for re-imaging (at the same imaging centre and using an imaging protocol identical to that for their first assessment) between Feb-Oct 2021. Their data provides a unique dataset comprehensively characterising brain and systemic health before and after SARS-CoV-2 infection that is linked to state-of-the-art clinical imaging. Here we provide a preliminary report of the CMR findings based on images available up to 22 Sep 2021.

## Methods

### Data

A longitudinal dataset consisting of 1,285 participants included in the UK Biobank SARS-CoV-2 D repeat imaging study, was used. These participants received CMR imaging scans before the start of the COVID-19 pandemic using a standard image acquisition protocol^14^. After the pandemic outbreak, participants with evidence of prior SARS-CoV-2 infection (n = 640) and a group of control participants (n = 645) matched for age, sex, ethnicity, imaging centre and date of first their first scan, were invited back for a repeated imaging scan using the same imaging protocols (**Figure S1**). Their infection status was determined from linkage to electronic health records (PCR antigen tests, primary care and hospital admissions data) and/or testing positive with two lateral flow antibody tests. The majority (95.3%) of cases were not hospitalised. Details about sample selection and case-control matching can be found elsewhere^15^. The two imaging scans were on average approximately 3.2 years apart. **Table 1** compares characteristics of the case and control groups (although matching was only partially complete at the time of preliminary data download).

**Table 1.** Characteristics of cases and controls at their first assessment visits. Weight, height, BMI, blood pressures, lifestyle and disease prevalence are reported using information at the first scan time, which was before the COVID pandemic outbreak. For categorical variables (e.g., sex, ethnicity, smoking, disease), the p-value is calculated using the chi-squared test. For continuous variables (e.g., age, weight, height), the nominal p-value is calculated using the two-sided student t-test. The Bonferroni threshold for multiple comparison (16 comparisons) is p_Bonf_ = 0.0031 for α = 0.05. BMI: body mass index; SBP: systolic blood pressure; DBP: diastolic blood pressure; COPD: chronic obstructive pulmonary disease.

|  | **Control**<br>**(n = 645)** | **Cases**<br>**(n = 640)** | **p value** |
| --- | --- | --- | --- |
| Sex (male/female) | 300 (46.5%) / 345 (53.5%) | 285 (44.5%) / 355 (55.5%) | 0.31 |
| Ethnicity (white/others) | 622 (96.4%) / 23 (3.6%) | 616 (96.2%) / 24 (3.8%) | 0.81 |
| Age at first imaging<br>assessment* (year) | 60.2 ± 7.4 | 59.4 ± 7.2 | 0.06 |
| Interval between scans (year) | 3.2 ± 1.6 | 3.2 ± 1.6 | 0.73 |
| Weight (kg) | 76.4 ± 15.1 | 77.2 ± 15.4 | 0.41 |
| Height (cm) | 169.5 ± 9.5 | 169.8 ± 9.0 | 0.67 |
| BMI (kg/m<sup>2</sup>) | 26.5 ± 4.5 | 26.7 ± 4.4 | 0.57 |
| SBP (mmHg) | 135.5 ± 17.5 | 134.4 ± 18.0 | 0.26 |
| DBP (mmHg) | 79.4 ± 9.8 | 79.3 ± 10.2 | 0.73 |
| Lifestyle |  |  |  |
| Current smoking | 19 (2.9%) | 28 (4.4%) | 0.08 |
| Self-reported diseases |  |  |  |
| Hypertension | 79 (12.2%) | 93 (14.5%) | 0.10 |
| Cardiac disease | 28 (4.3%) | 28 (4.4%) | 0.96 |
| Diabetes | 16 (2.5%) | 23 (3.6%) | 0.13 |
| Asthma | 56 (8.7%) | 51 (8.0%) | 0.50 |
| COPD | 1 (0.2%) | 1 (0.2%) | 0.99 |
| Stroke | 4 (0.6%) | 3 (0.5%) | 0.57 |

\*Before COVID-19 for cases.

### Image analysis

Cardiac and aortic imaging phenotypes were extracted from the CMR images using an automated cardiac image analysis pipeline^16–18^. Each phenotype describes one aspect of structure or function for the four cardiac chambers (left ventricle: LV; right ventricle: RV; left atrium: LA; right atrium: RA) or the two aortic cross-sections (ascending aorta: AAo; descending aorta: DAo). Convolutional neural networks were used for segmentation of cardiac or aortic cine images^16–18^, from which volumes or areas of the cardiac or aortic structures were calculated. The ejection fractions of the cardiac chambers were calculated from the maximal and minimal volumes across the cardiac cycle. The aortic distensibilities were calculated from the maximal, minimal cross-sectional areas of the aorta and central pulse pressure^14^. Global circumferential, radial and longitudinal strains of the LV myocardium were evaluated by use of non-rigid image registration techniques for cardiac motion tracking^3^. In total, we utilised 27 cardiac and aortic imaging measures for subsequent statistical analyses. **Table 2** reports these measures at the first assessments for the case and control groups and confirms the comparability of CMR measures between two groups at the baseline.

**Table 2.** Comparison of cardiac imaging phenotypes patients at the first assessment visit between cases and controls. The nominal p-value is reported using the two-sided student T-test. None of the group differences were statistically significant. The Bonferroni threshold for multiple comparison (27 imaging phenotypes) is p_Bonf_ = 0.0019 for α = 0.05.

|  | Control<br>(n = 645) | Cases<br>(n = 640) | p<br>value |
| --- | --- | --- | --- |
| **Left ventricle** |  |  |  |
| LV end-diastolic volume (mL) | 150.3 ± 31.7 | 149.9 ± 32.8 | 0.86 |
| LV end-systolic volume (mL) | 61.6 ± 17.4 | 61.9 ± 17.5 | 0.79 |
| LV stroke volume (mL) | 88.6 ± 18.6 | 88.0 ± 19.2 | 0.58 |
| LV ejection fraction (%) | 59.3 ± 5.5 | 59.0 ± 5.6 | 0.32 |
| LV cardiac output (L/min) | 5.4 ± 1.2 | 5.4 ± 1.3 | 0.80 |
| LV myocardial mass (g) | 86.1 ± 21.4 | 86.7 ± 22.8 | 0.63 |
| Global peak strain Ecc (%) | 22.4 ± 3.1 | 22.0 ± 3.1 | 0.05 |
| Global peak strain Err (%) | 44.3 ± 7.4 | 44.0 ± 7.9 | 0.55 |
| Global peak strain Ell (%) | 18.5 ± 2.7 | 18.5 ± 2.6 | 0.77 |
| **Right ventricle** |  |  |  |
| RV end-diastolic volume (mL) | 159.6 ± 36.1 | 159.0 ± 37.4 | 0.79 |
| RV end-systolic volume (mL) | 69.2 ± 20.8 | 69.5 ± 21.3 | 0.77 |
| RV stroke volume (mL) | 90.4 ± 19.7 | 89.5 ± 20.2 | 0.43 |
| RV ejection fraction (%) | 57.0 ± 5.8 | 56.7 ± 5.9 | 0.26 |
| **Left atrium** |  |  |  |
| LA maximal volume (mL) | 73.2 ± 21.0 | 73.1 ± 21.5 | 0.92 |
| LA minimal volume (mL) | 29.0 ± 12.4 | 29.1 ± 13.3 | 0.86 |
| LA stroke volume (mL) | 44.2 ± 11.5 | 44.0 ± 11.5 | 0.70 |
| LA ejection fraction (%) | 61.4 ± 8.3 | 61.4 ± 8.6 | 0.90 |
| **Right atrium** |  |  |  |
| RA maximal volume (mL) | 87.2 ± 26.5 | 85.9 ± 26.6 | 0.41 |
| RA minimal volume (mL) | 47.3 ± 18.4 | 46.0 ± 17.6 | 0.20 |
| RA stroke volume (mL) | 39.8 ± 12.7 | 39.9 ± 13.1 | 0.94 |
| RA ejection fraction (%) | 46.3 ± 9.0 | 46.9 ± 8.4 | 0.25 |
| **Ascending aorta** |  |  |  |
| AAo maximal area (mm<sup>2</sup>) | 838.0 ± 191.7 | 813.9 ± 184.5 | 0.03 |
| AAo minimal area (mm<sup>2</sup>) | 751.1 ± 186.5 | 729.1 ± 179.6 | 0.05 |
| AAo distensibility (10<sup>-3</sup> mmHg<sup>-1</sup>) | 2.2 ± 1.2 | 2.3 ± 1.5 | 0.13 |
| **Descending aorta** |  |  |  |
| DAo maximal area (mm<sup>2</sup>) | 465.0 ± 99.4 | 461.5 ± 99.6 | 0.56 |
| DAo minimal area (mm<sup>2</sup>) | 403.9 ± 93.9 | 401.8 ± 95.1 | 0.71 |
| DAo distensibility (10<sup>-3</sup> mmHg<sup>-1</sup>) | 2.7 ± 1.3 | 2.8 ± 1.5 | 0.36 |

### Statistical analysis

Here we focused on analysing the longitudinal changes of the cardiac and aortic imaging measures. We assumed a multiple linear regression model for each imaging measure,

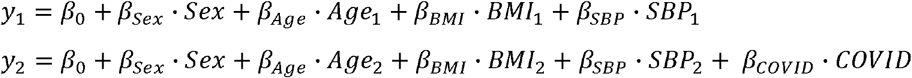

where *y* denotes an imaging measure, the subscripts 1 and 2 denote time point 1 and time point 2, *β* denotes the coefficient or the effect size of each factor. We included Sex, Age, BMI and SBP into the regression model, where are common risk factors that influence the heart and aorta. Since the first imaging scan was taken before the COVID pandemic, we did not include the COVID factor for. Therefore, the longitudinal change of a phenotype can be calculated by,

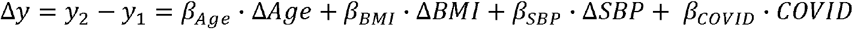

In case the change Δ is also influenced by Sex, the final regression model we adopted (**Model 1**) was,

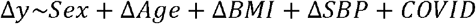

We also fitted a more complex model (**Model 2**) to account for the influences of more factors, which was formulated as,

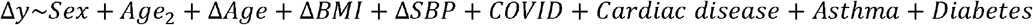

where *Age*_*2*_ denotes the age at the second imaging scan, which was included as the effect of COVID was shown to increase dramatically with age^19,20^. Cardiac disease, asthma and diabetes were common co-morbid conditions in hospitalised COVID patients, as shown in Table 2. These diseases were defined using the self-reported disease code^16^.

## Results

### Lack of CMR evidence for cardiac structural and functional differences between cases and controls

At baseline, prior to the COVID-19 pandemic, participants in the case and control groups had similar cardiac CMR measures (**Table 2**).

We then tested for differences in longitudinal changes in the cardiac and aortic imaging phenotypes between cases and controls. No significant differences (after correction for multiple comparisons) in changes were found for the cases relative to controls (**Figure 1, Table 3**).

**Table 3.** Comparison of the longitudinal changes of cardiac imaging measures between cases and controls. Columns 2 and 3 report the longitudinal changes between two imaging scans. Column 4 reports the p-value of the two-sided student T-test. The Bonferroni threshold for multiple comparison (27 imaging phenotypes) is p_Bonf_ = 0.0019 for α= 0.05.

|  | **Control**<br>**(n = 645)** | **Cases**<br>**(n = 640)** | **p**<br>**value** |
| --- | --- | --- | --- |
| **Left ventricle** |  |  |  |
| LV end-diastolic volume (mL) | -1.8 ± 14.0 | -1.8 ± 14.8 | 1.00 |
| LV end-systolic volume (mL) | -0.4 ± 9.2 | 0.1 ± 9.3 | 0.36 |
| LV stroke volume (mL) | -1.4 ± 12.6 | -1.9 ± 12.7 | 0.50 |
| LV ejection fraction (%) | -0.1 ± 5.3 | -0.4 ± 5.2 | 0.25 |
| LV cardiac output (L/min) | -0.3 ± 1.0 | -0.3 ± 1.0 | 0.60 |
| LV myocardial mass (g) | 0.1 ± 6.6 | 0.2 ± 6.3 | 0.79 |
| Global peak strain Ecc (%) | -0.2 ± 2.5 | -0.1 ± 2.3 | 0.90 |
| Global peak strain Err (%) | 0.3 ± 6.7 | 0.1 ± 6.7 | 0.71 |
| Global peak strain Ell (%) | -0.2 ± 2.6 | -0.3 ± 2.4 | 0.59 |
| **Right ventricle** |  |  |  |
| RV end-diastolic volume (mL) | -1.8 ± 15.4 | -0.8 ± 15.6 | 0.23 |
| RV end-systolic volume (mL) | -0.1 ± 8.9 | 0.1 ± 8.7 | 0.61 |
| RV stroke volume (mL) | -1.7 ± 13.1 | -0.9 ± 13.2 | 0.29 |
| RV ejection fraction (%) | -0.4 ± 4.9 | -0.3 ± 4.9 | 0.63 |
| **Left atrium** |  |  |  |
| LA maximal volume (mL) | -1.7 ± 14.4 | -0.7 ± 15.7 | 0.21 |
| LA minimal volume (mL) | -0.3 ± 8.8 | 0.8 ± 11.2 | 0.06 |
| LA stroke volume (mL) | -1.5 ± 8.6 | -1.5 ± 8.9 | 0.94 |
| LA ejection fraction (%) | -0.4 ± 7.8 | -1.3 ± 9.1 | 0.05 |
| **Right atrium** |  |  |  |
| RA maximal volume (mL) | -0.2 ± 16.3 | -0.3 ± 18.1 | 0.91 |
| RA minimal volume (mL) | 0.1 ± 11.6 | 1.3 ± 11.9 | 0.09 |
| RA stroke volume (mL) | -0.3 ± 11.6 | -1.5 ± 11.1 | 0.05 |
| RA ejection fraction (%) | -0.3 ± 8.6 | -1.3 ± 8.6 | 0.03 |
| **Ascending aorta** |  |  |  |
| AAo maximal area (mm<sup>2</sup>) | 21.9 ± 45.1 | 30.0 ± 54.3 | 0.09 |
| AAo minimal area (mm<sup>2</sup>) | 24.8 ± 46.8 | 34.0 ± 51.3 | 0.05 |
| AAo distensibility (10<sup>-3</sup> mmHg<sup>-1</sup>) | -0.2 ± 1.1 | -0.3 ± 1.2 | 0.36 |
| **Descending aorta** |  |  |  |
| DAo maximal area (mm<sup>2</sup>) | 9.7 ± 27.2 | 12.3 ± 29.1 | 0.34 |
| DAo minimal area (mm<sup>2</sup>) | 13.5 ± 26.5 | 14.9 ± 28.5 | 0.61 |
| DAo distensibility (10<sup>-3</sup> mmHg<sup>-1</sup>) | -0.3 ± 1.2 | -0.3 ± 1.4 | 1.00 |

**Figure 1.**
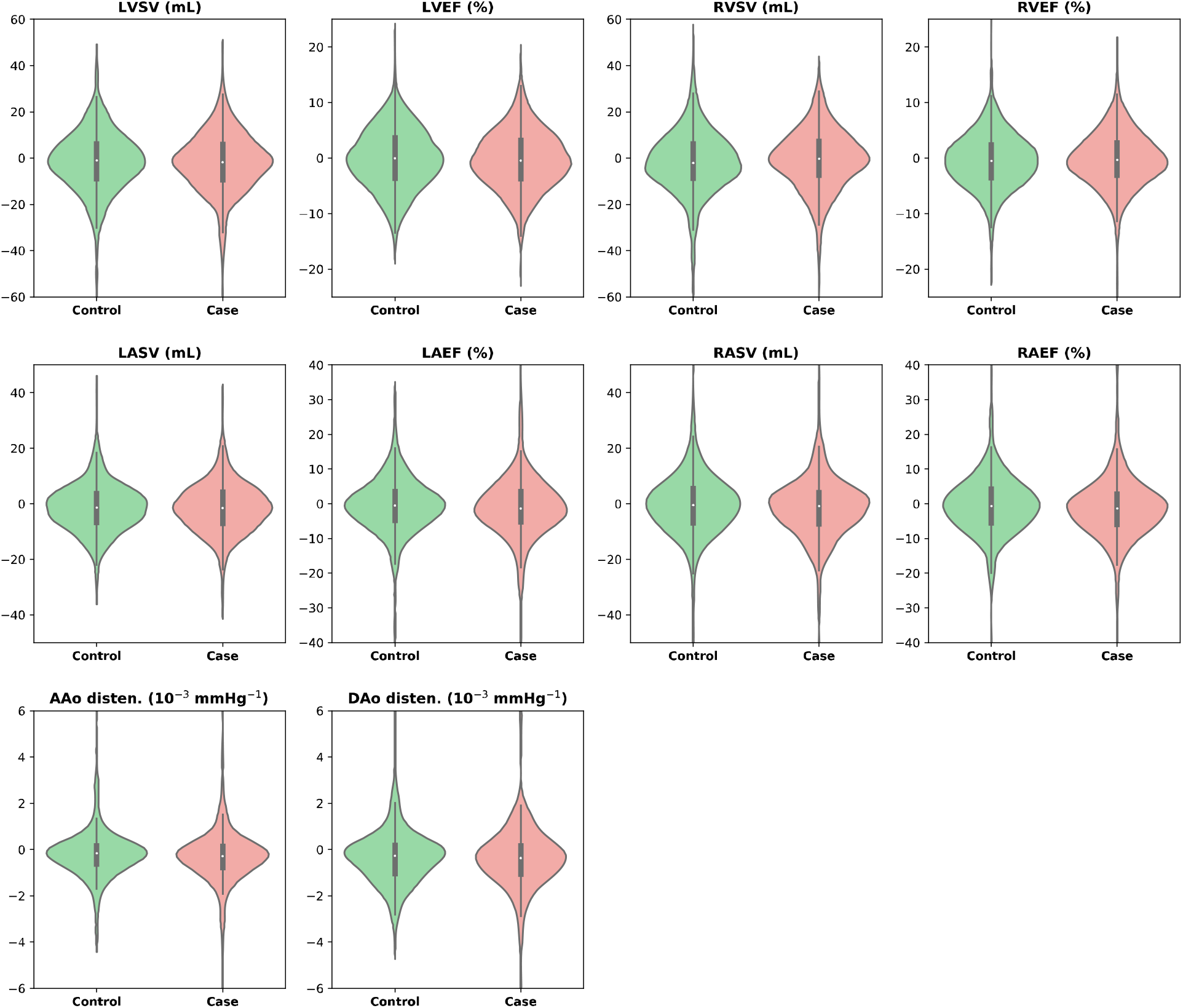
Comparison of the longitudinal changes of a selected of imaging phenotypes for the four cardiac chambers and two aortic cross-sections between cases and controls. LVSV: left ventricular stroke volume; LVEF: left ventricular ejection fraction; RVSV: right ventricular stroke volume; RVEF: right ventricular ejection fraction; LASV: left atrial stroke volume; LAEF: left atrial ejection fraction; RASV: right atrial stroke volume; RAEF: right atrial ejection fraction; AAo disten.: ascending aorta distensibility; DAo disten.: descending aorta distensibility

Although cases and controls were approximately matched, direct comparisons of the measures between the two groups have potential confounds arising from differences that could not be matched between the groups. We applied linear regression models including those confounds that were not matched to better estimate the independent effects of COVID-19 on longitudinal changes. Neither a simpler (Model 1) or more complex (Model 2) showed significant longitudinal differences of cardiac structure and function between cases and controls (**Table 4, Figure 2**).

**Table 4.**
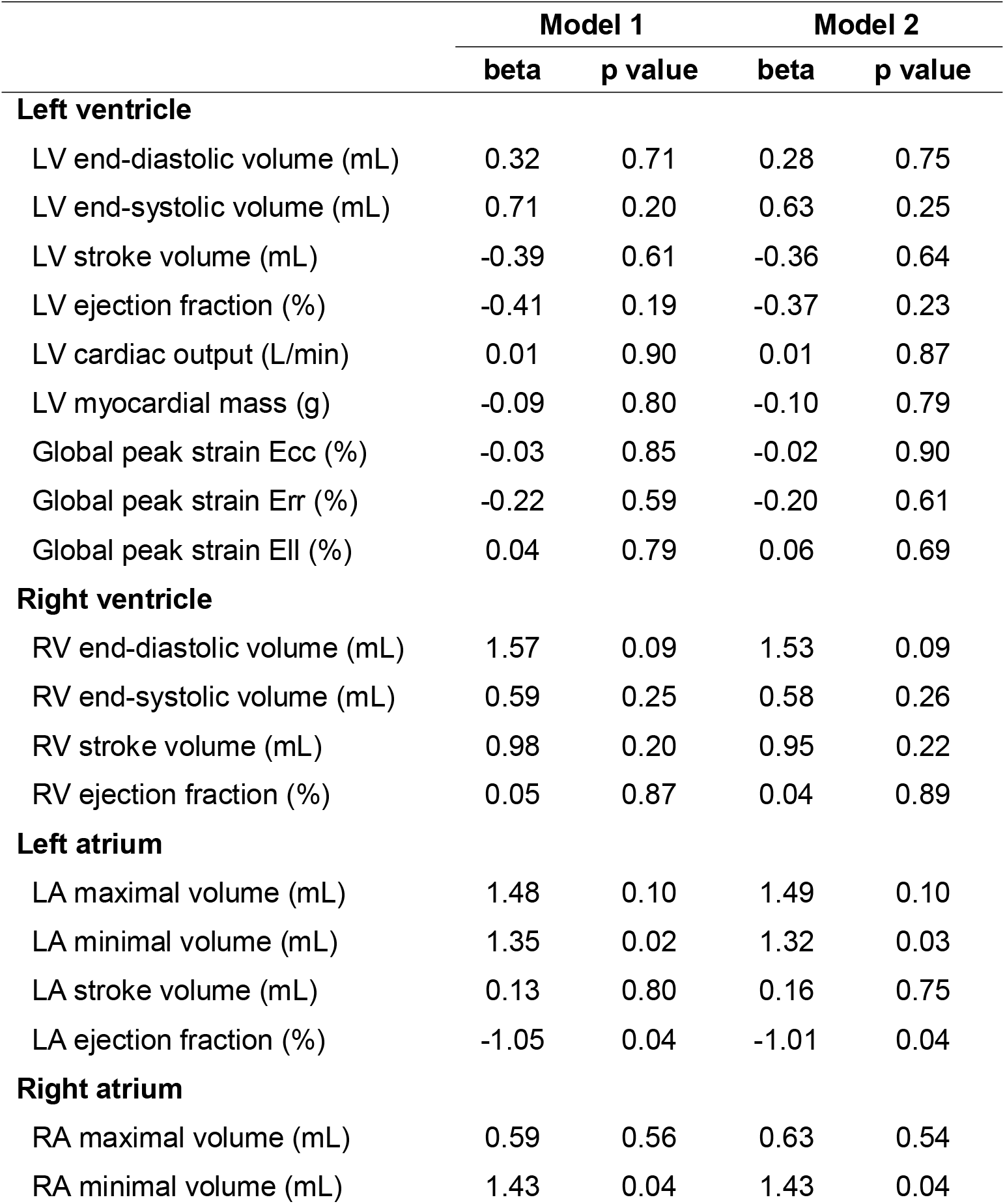
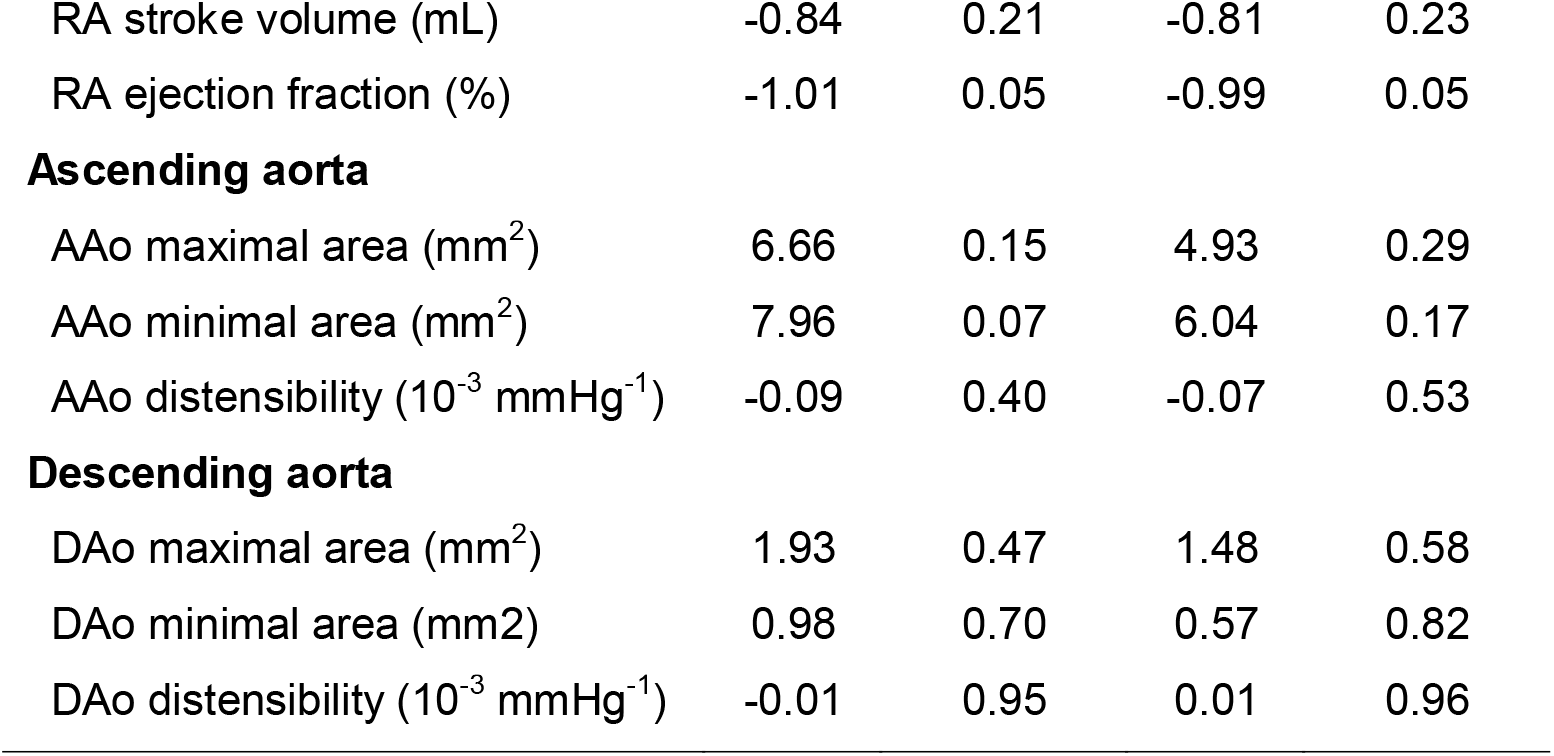
Regression modelling of longitudinal case-control differences in cardiac imaging phenotypes. Model 1: ΔIDP ∼ Sex + ΔAge + ΔBMI + ΔSBP + COVID; Model 2: ΔIDP ∼ Sex + Age_2_ + ΔAge + ΔBMI + ΔSBP + Cardiac disease + Asthma + Diabetes + COVID. Regression analyses were applied to both controls and COVID cases. The columns report the regression coefficients and the p-values for COVID. Nominal p-values are reported. The Bonferroni threshold for multiple comparison (27 imaging phenotypes) is p_Bonf_ = 0.0019 for α = 0.05.

|  | Model 1 |  | Model 2 |  |
| --- | --- | --- | --- | --- |
|  | beta | p value | beta | p value |
| **Left ventricle** |  |  |  |  |
| LV end-diastolic volume (mL) | 0.32 | 0.71 | 0.28 | 0.75 |
| LV end-systolic volume (mL) | 0.71 | 0.20 | 0.63 | 0.25 |
| LV stroke volume (mL) | -0.39 | 0.61 | -0.36 | 0.64 |
| LV ejection fraction (%) | -0.41 | 0.19 | -0.37 | 0.23 |
| LV cardiac output (L/min) | 0.01 | 0.90 | 0.01 | 0.87 |
| LV myocardial mass (g) | -0.09 | 0.80 | -0.10 | 0.79 |
| Global peak strain Ecc (%) | -0.03 | 0.85 | -0.02 | 0.90 |
| Global peak strain Err (%) | -0.22 | 0.59 | -0.20 | 0.61 |
| Global peak strain Ell (%) | 0.04 | 0.79 | 0.06 | 0.69 |
| **Right ventricle** |  |  |  |  |
| RV end-diastolic volume (mL) | 1.57 | 0.09 | 1.53 | 0.09 |
| RV end-systolic volume (mL) | 0.59 | 0.25 | 0.58 | 0.26 |
| RV stroke volume (mL) | 0.98 | 0.20 | 0.95 | 0.22 |
| RV ejection fraction (%) | 0.05 | 0.87 | 0.04 | 0.89 |
| **Left atrium** |  |  |  |  |
| LA maximal volume (mL) | 1.48 | 0.10 | 1.49 | 0.10 |
| LA minimal volume (mL) | 1.35 | 0.02 | 1.32 | 0.03 |
| LA stroke volume (mL) | 0.13 | 0.80 | 0.16 | 0.75 |
| LA ejection fraction (%) | -1.05 | 0.04 | -1.01 | 0.04 |
| **Right atrium** |  |  |  |  |
| RA maximal volume (mL) | 0.59 | 0.56 | 0.63 | 0.54 |
| RA minimal volume (mL) | 1.43 | 0.04 | 1.43 | 0.04 |
| RA stroke volume (mL) | -0.84 | 0.21 | -0.81 | 0.23 |
| RA ejection fraction (%) | -1.01 | 0.05 | -0.99 | 0.05 |
| Ascending aorta |  |  |  |  |
| AAo maximal area (mm<sup>2</sup>) | 6.66 | 0.15 | 4.93 | 0.29 |
| AAo minimal area (mm<sup>2</sup>) | 7.96 | 0.07 | 6.04 | 0.17 |
| AAo distensibility (10<sup>-3</sup> mmHg<sup>-1</sup>) | -0.09 | 0.40 | -0.07 | 0.53 |

**Table 4. Regression modelling of longitudinal case-control differences in cardiac imaging phenotypes.**
|  |  |  |  |  |
| --- | --- | --- | --- | --- |
| DAo maximal area (mm<sup>2</sup>) | 1.93 | 0.47 | 1.48 | 0.58 |
| DAo minimal area (mm<sup>2</sup>) | 0.98 | 0.70 | 0.57 | 0.82 |
| DAo distensibility (10<sup>-3</sup> mmHg<sup>-1</sup>) | -0.01 | 0.95 | 0.01 | 0.96 |

**Figure 2.**
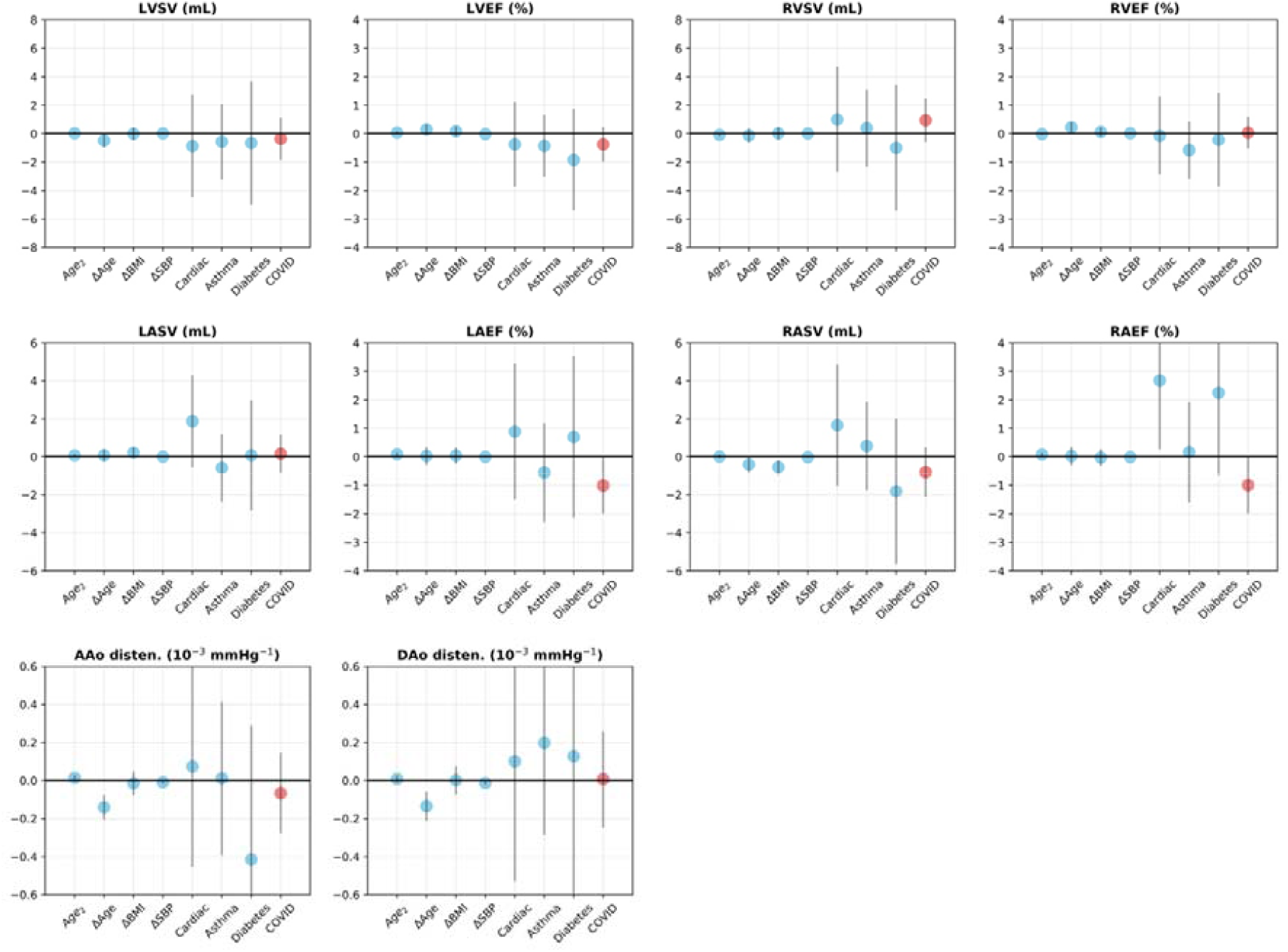
Plots of coefficients for regression Model 2 comparing cases and controls. The regression coefficient associated with SARS-COV-2 infection (coloured red) is described as the “COVID factor” (with directionality for all coefficients described as difference that for scan 2 relative to that for scan 1, e.g., a negative value indicates a reduction in the measure between the two scans). The label “Cardiac” denotes prior cardiac disease. The gray bars denote the 95% confidence interval. For each factor, the coefficient describes the effect size with a change in the variable by 1 unit. For example, Δ*Age* increases by 1 year or COVID status changes from 0 to 1.

### Post-hoc exploration of differences between hospitalised and non-hospitalised cases at first assessment and longitudinally

Previous CMR investigations of people ascertained and imaged only *after* diagnosis with COVID-19 highlighted a higher frequency of cardiac structural or functional abnormalities than expected in a healthy population^5,6^. To explore the hypothesis that this could reflect a greater susceptibility to more severe COVID-19 disease amongst people with pre-existing CMR findings, we compared cases who were hospitalised relative to those who were not. Thirty of the cases had been hospitalised for COVID-19 as identified by the ICD-10 diagnosis codes U07.1 or U07.2 available from the UK Biobank inpatient data. **Table S1** compares the participant characteristics of hospitalized and non-hospitalized cases.

Compared with cases who were *not* hospitalised, cases (n = 30) who were hospitalised had a larger mean descending aorta minimal area at their first imaging assessment (**Table S2**). No significant longitudinal differences between the two sub-groups of cases were found (**Figure S2, Table S3**). However, the numbers are small and the statistical power to detect differences thus is low. We applied both Model 1 and Model 2 in exploratory analyses of longitudinal changes in CMR measures between hospitalised and non-hospitalised cases by replacing the COVID covariate by a hospitalisation covariate expressed as a binary variable. Regression modelling of longitudinal changes showed an increase in RA stroke volume for the hospitalised cases **(Table S4)**.

## Discussion

There is considerable interest in understanding possible cardiac complications of COVID-19 because of the still growing numbers of people infected with SARS-CoV-2 globally. There has been reason to believe that cardiac involvement could be clinically significant. Many survivors of COVID-19 experience symptoms potentially referable to the heart, as well as non-specific long-term symptoms including fatigue and increased breathlessness^21^. Multiple case studies of people studied after COVID-19 have suggested that symptoms might arise from infection-associated injury to the heart: a systematic literature search found evidence for one or more abnormal CMR findings in nearly half of recovered COVID-19 patients reported at the time^22^.

However, in marked contrast to this case report literature, our results do not suggest that CMR abnormalities are either common after infection or likely to be independently clinically significant. Our data are unique, as the longitudinal design of the UK Biobank study with images from both before and after SARS-CoV-2 infection largely removes the confound of “reverse causation bias”. We did not observe any significant differences in cardiac CMR measures after infection in the cases compared to the matched non-infected control group. However, the majority of the infections were mild. Other recent data also have suggested that cardiac abnormalities are infrequent after milder, predominantly community-based SARS-CoV-2 infections. For example, a study of 149 health care workers assessed using CMR imaging 6 months after mild or asymptomatic SARS-CoV-2 infections concluded that cardiovascular abnormalities are no more common in people after infection than in the general population^23^.

We hypothesise that the discrepancy between our negative observations with a prospective case-control design comparing changes in CMR imaging from *before and after* SARS-CoV-2 infection and the high frequency of abnormalities suggested by earlier case studies arises because the latter were based on examinations of images acquired only *after* infection (which, in most of the published cases has been severe enough to need hospitalisation). The high incidence of abnormalities in earlier case studies may reflect both reporting bias and confounding by “reverse causation”. While the numbers are small, our *post hoc* exploration of hospitalised vs. non-hospitalised cases provides some evidence that baseline characteristics may determine susceptibility or the course of infection. An earlier preliminary report of relationships between CMR imaging measures from the UK Biobank volunteers at first imaging assessments and likelihood of subsequent SARS-CoV-2 infection also suggested that people with adverse CMR phenotypes have a higher likelihood of SARS-CoV-2 infection, independent of classical cardiovascular risk factors^24^. Other possible explanations of our largely negative findings are that the cases were predominantly milder (not involving hospitalisations) or that CMR abnormalities after infection may be reversible in part (and thus dependent on timing of observations with respect to the acute disease), e.g., lower ventricular circumferential or radial strain measures for COVID patients < 8 weeks after recovery from the acute infection than those for > 8 weeks was reported recently^25^. Similarly, right ventricular function and myocardial tissue characteristics were seen to recover in another study of hospitalised patients from three to six months after acute infection^26^. Further modelling to explore the possible dependence of CMR case-control differences on time since resolution of acute infections is needed to explore this possibility.

A limitation of our study is that all cardiac and aortic measurements were performed using fully automated image segmentation, although the method was validated previously using UK Biobank data^16,17^. The population studied also is limited to volunteers from amongst UK Biobank participants. The extent to which findings can be generalised particularly to more severe cases of COVID-19 is unknown. As noted above, the exploratory comparison between hospitalised and non-hospitalised infections is based on a small number of hospitalised cases, although the numbers are similar to those in some of the previous case series reporting abnormalities after COVID-19^4^. Finally, the analyses have only considered structural and functional CMR measures. Additional analyses ongoing that will incorporate other cardiac functional data (e.g., ECG) and more detailed examinations of cardiac phenotypes (*SEP, unpublished current investigations*) could identify longitudinal case-control differences not detected here.

Nonetheless, our observations suggest that persistent, clinically significant cardiac complications of SARS-CoV-2 infection are not common in the middle-aged to older UK participants in this UK Biobank pre- and post-infection imaging study.

## Data Availability

The automated cardiac and aortic segementation data generated for this study has been deposited with UK Biobank for access by other researchers on application to UK Biobank (https://www.ukbiobank.ac.uk/).

## Acknowledgments

PMM acknowledges generous personal and research support from the Edmond J Safra Foundation and Lily Safra, a National Institute for Health Research (NIHR) Senior Investigator Award, the UK Dementia Research Institute and the NIHR Biomedical Research Centre at Imperial College London. WB acknowledges support from EPSRC SmartHeart Programme (EP/P001009/1). SEP acknowledges support from the National Institute for Health Research Barts Biomedical Research Centre. NA and ZRE acknowledge support from the NIHR through Academic Clinical Lecturer funding. ZRE was supported by British Heart Foundation Clinical Research Training Fellowship No. FS/17/81/33318. BR and SN acknowledge support from the Oxford NIHR Biomedical Research Centre and the British Heart Foundation. BR is also supported by the British Heart Foundation Oxford Centre of Research Excellence Transition Clinical Intermediate Fellowship (RE/18/3/34214). NCH acknowledges support from the UK Medical Research Council (MRC MC_UU_12011/1), and NIHR Southampton Biomedical Research Centre, University of Southampton, and University Hospital Southampton. RC Is a British Heart Foundation Chairholder.

## Supplementary Figures

**Figure S1.**
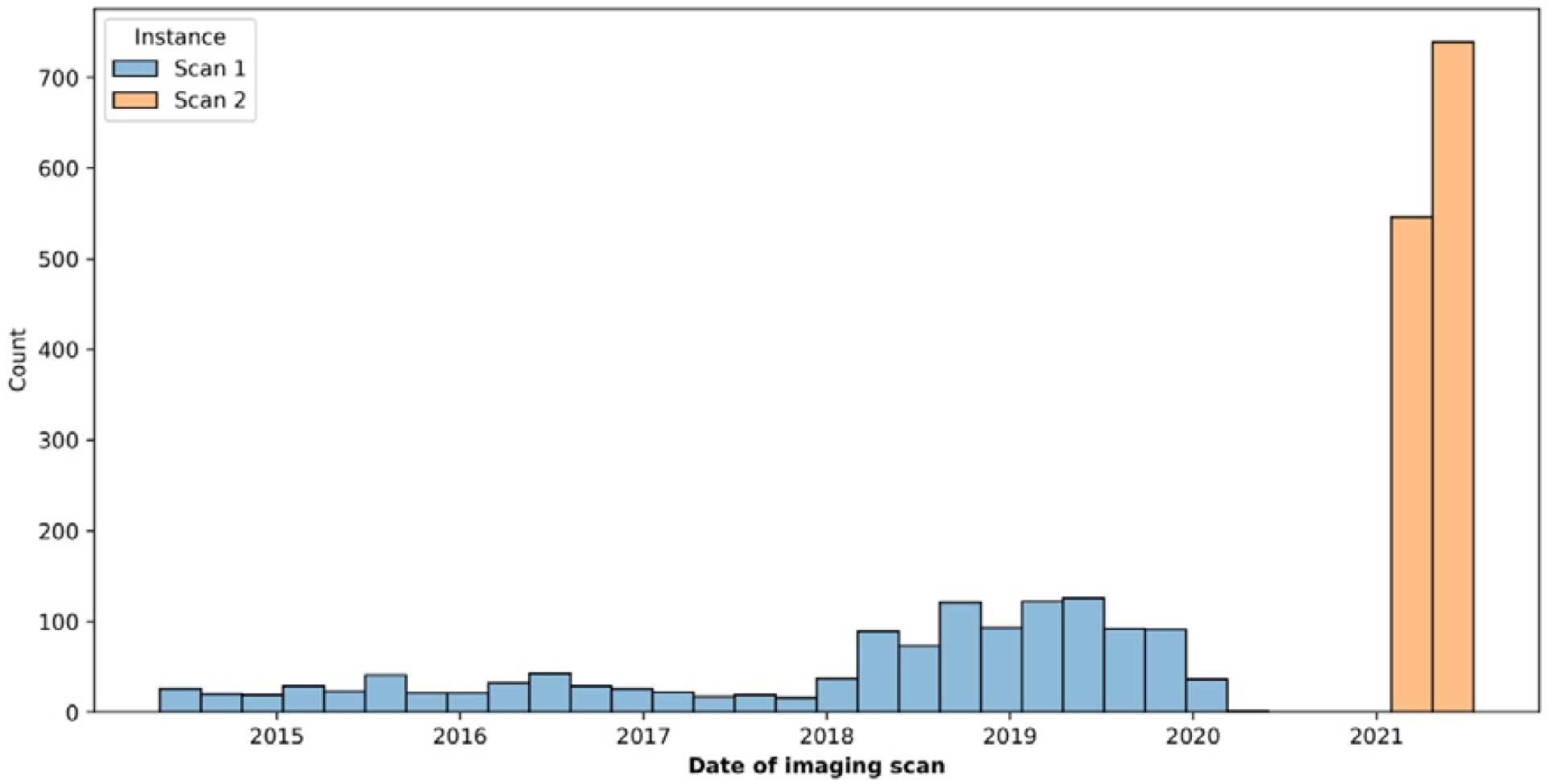
Timeline of the repeat CMR imaging scans for 1,285 UK Biobank participants. The first imaging scans (blue) were acquired between May 2014 and March 2021. The second imaging scans (orange) were acquired between February 2021 and July 2021.

**Figure S2.**
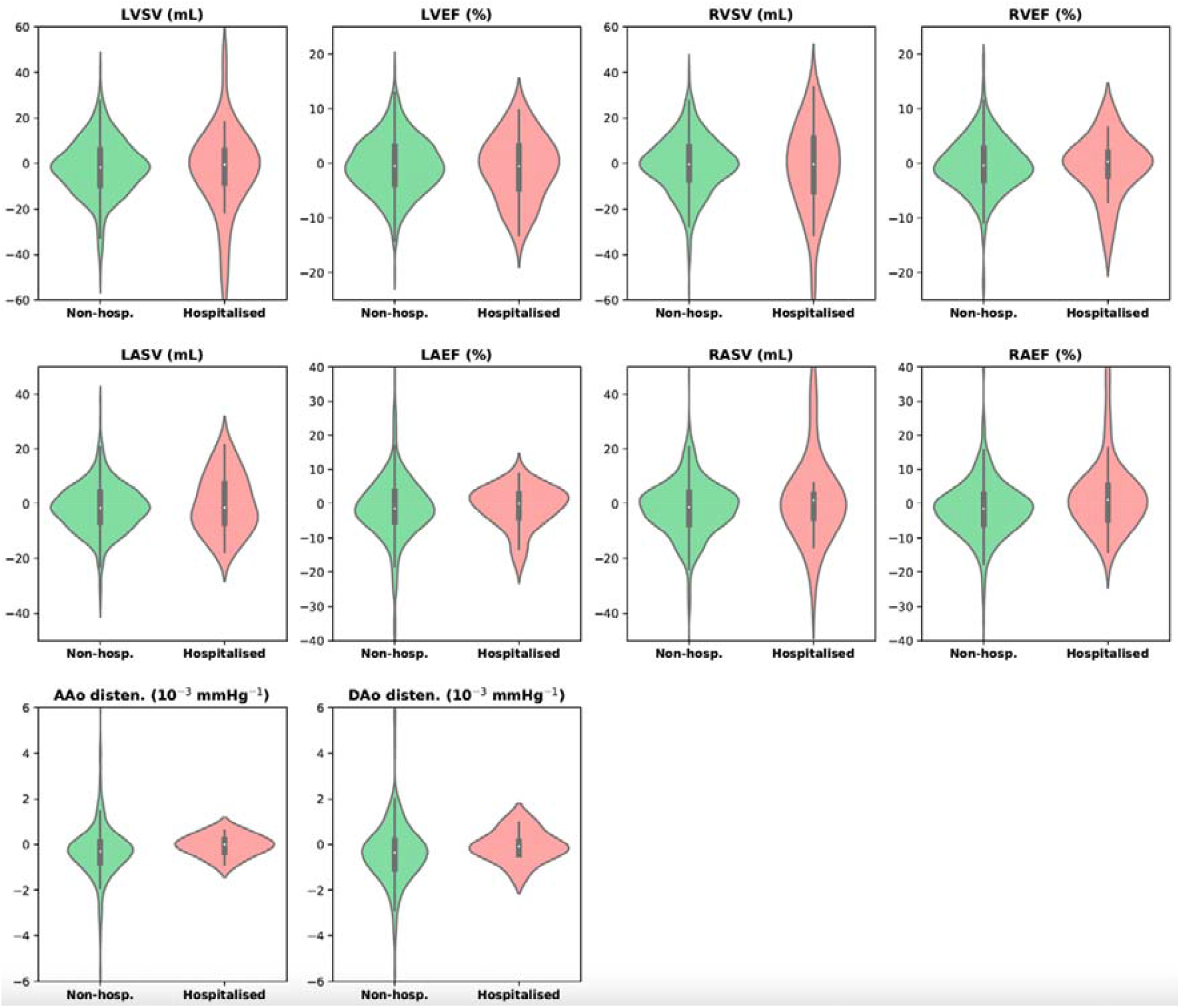
Comparison of the longitudinal changes of a selected of imaging phenotypes for the four cardiac chambers and two aortic cross-sections between hospitalised and non-hospitalised cases.

## Supplementary Tables

**Table S1.**
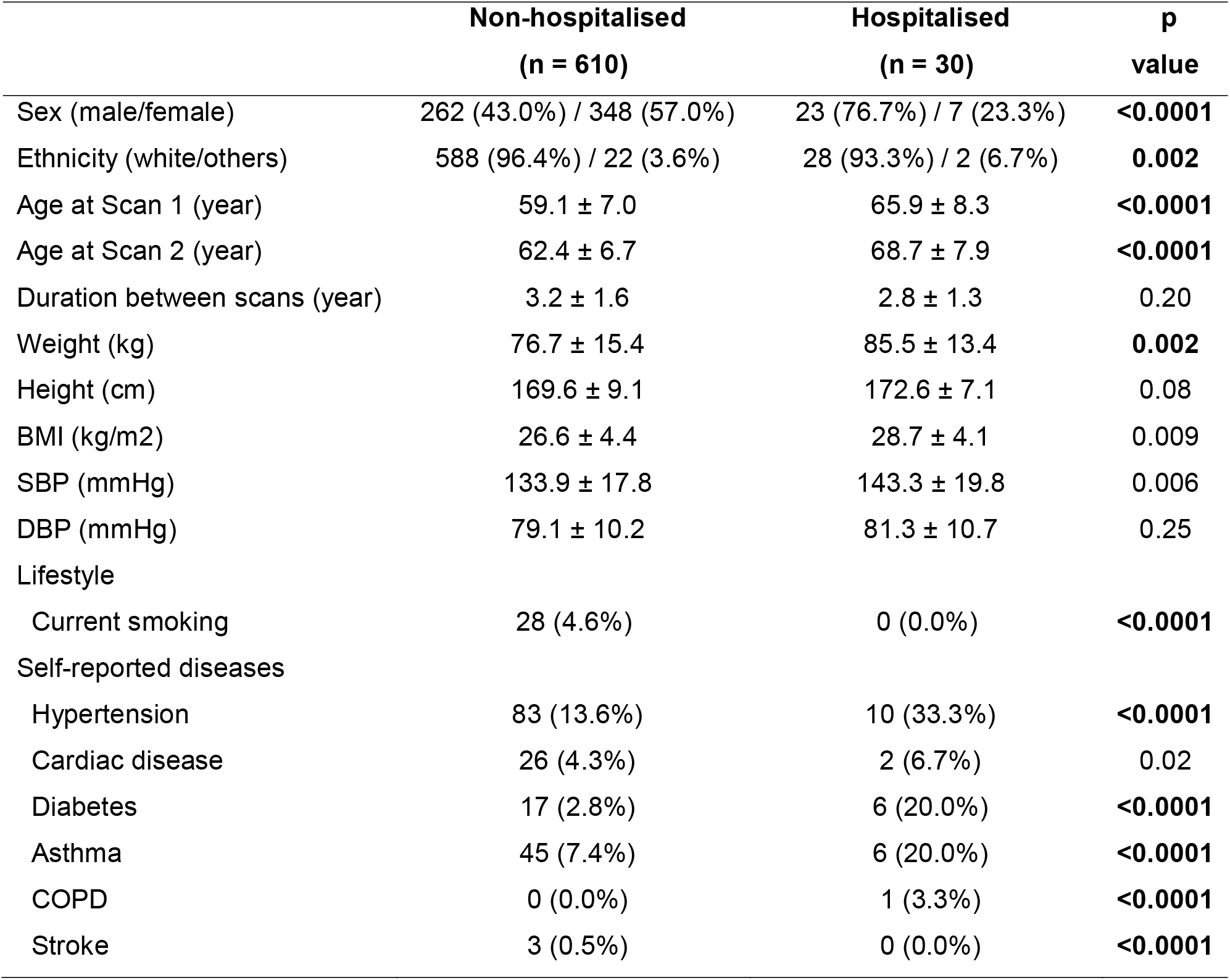
Comparison of participant characteristics between non-hospitalised and hospitalised SARS-CoV-2 cases. Hospitalisation was determined using the linked ICD10 code for COVID-19 (U07.1 or U07.2). The Bonferroni threshold for multiple comparison (16 comparisons) is p_Bonf_ = 0.0031 for α = 0.05. Bold font indicates a p-value < p_Bonf_.

**Table S2.** Comparison of cardiac imaging phenotypes at the first scan between hospitalised and non-hospitalised cases. The p-value is reported using the two-sided student T-test. The Bonferroni threshold for multiple comparison (27 imaging phenotypes) is p_Bonf_ = 0.0019 for α = 0.05. Bold font indicates a p-value < p_Bonf_.

|  | Non-hospitalised<br>(n = 610) | Hospitalised<br>(n = 30) | p<br>value |
| --- | --- | --- | --- |
| **Left ventricle** |  |  |  |
| LV end-diastolic volume (mL) | 149.5 ± 32.6 | 158.2 ± 35.1 | 0.16 |
| LV end-systolic volume (mL) | 61.6 ± 17.2 | 68.2 ± 23.4 | 0.04 |
| LV stroke volume (mL) | 87.9 ± 19.2 | 90.0 ± 20.0 | 0.57 |
| LV ejection fraction (%) | 59.0 ± 5.5 | 57.3 ± 7.0 | 0.10 |
| LV cardiac output (L/min) | 5.4 ± 1.3 | 5.7 ± 1.6 | 0.19 |
| LV myocardial mass (g) | 86.3 ± 22.7 | 95.1 ± 23.9 | 0.04 |
| Global peak strain Ecc (%) | 22.1 ± 3.1 | 20.9 ± 3.7 | 0.05 |
| Global peak strain Err (%) | 44.1 ± 7.9 | 41.8 ± 7.6 | 0.13 |
| Global peak strain Ell (%) | 18.5 ± 2.6 | 18.5 ± 2.5 | 0.93 |
| **Right ventricle** |  |  |  |
| RV end-diastolic volume (mL) | 158.5 ± 37.4 | 169.3 ± 35.9 | 0.12 |
| RV end-systolic volume (mL) | 69.1 ± 21.2 | 77.1 ± 22.2 | 0.04 |
| RV stroke volume (mL) | 89.4 ± 20.2 | 92.2 ± 20.8 | 0.46 |
| RV ejection fraction (%) | 56.8 ± 5.8 | 54.6 ± 7.1 | 0.05 |
| **Left atrium** |  |  |  |
| LA maximal volume (mL) | 73.0 ± 21.6 | 75.1 ± 20.9 | 0.59 |
| LA minimal volume (mL) | 29.0 ± 13.3 | 30.5 ± 11.9 | 0.56 |
| LA stroke volume (mL) | 43.9 ± 11.5 | 44.6 ± 12.0 | 0.74 |
| LA ejection fraction (%) | 61.4 ± 8.6 | 60.2 ± 8.2 | 0.47 |
| **Right atrium** |  |  |  |
| RA maximal volume (mL) | 85.7 ± 26.7 | 90.0 ± 25.0 | 0.39 |
| RA minimal volume (mL) | 45.8 ± 17.7 | 49.9 ± 15.3 | 0.21 |
| RA stroke volume (mL) | 39.9 ± 13.0 | 40.1 ± 14.4 | 0.93 |
| RA ejection fraction (%) | 47.0 ± 8.4 | 44.1 ± 8.9 | 0.07 |
| **Ascending aorta** |  |  |  |
| AAo maximal area (mm<sup>2</sup>) | 811.6 ± 184.5 | 865.7 ± 179.0 | 0.16 |
| AAo minimal area (mm<sup>2</sup>) | 726.4 ± 179.5 | 788.1 ± 175.1 | 0.10 |
| AAo distensibility (10<sup>-3</sup> mmHg<sup>-1</sup>) | 2.3 ± 1.6 | 1.8 ± 1.1 | 0.12 |
| **Descending aorta** |  |  |  |
| DAo maximal area (mm<sup>2</sup>) | 459.0 ± 97.7 | 517.2 ± 124.1 | 0.01 |
| DAo minimal area (mm<sup>2</sup>) | 399.0 ± 93.2 | 461.5 ± 117.0 | **0.0016** |
| DAo distensibility (10<sup>-3</sup> mmHg<sup>-1</sup>) | 2.9 ± 1.5 | 2.1 ± 0.9 | 0.02 |

**Table S3.** Comparison of the longitudinal changes of cardiac imaging phenotypes between hospitalised and non-hospitalised cases. Columns 2 and 3 report the longitudinal changes between two imaging scans. Column 4 reports the p-value of the two-sided student T-test. The Bonferroni threshold for multiple comparison (27 imaging phenotypes) is p_Bonf_ = 0.0019 for α = 0.05. Bold font indicates a p-value < p_Bonf_.

|  | Non-hospitalised<br>(n = 610) | Hospitalised<br>(n = 30) | p<br>value |
| --- | --- | --- | --- |
| **Left ventricle** |  |  |  |
| LV end-diastolic volume (mL) | -1.8 ± 14.1 | -1.9 ± 26.4 | 0.97 |
| LV end-systolic volume (mL) | 0.0 ± 9.0 | 1.1 ± 14.8 | 0.55 |
| LV stroke volume (mL) | -1.8 ± 12.4 | -3.0 ± 18.3 | 0.63 |
| LV ejection fraction (%) | -0.4 ± 5.2 | -0.8 ± 5.8 | 0.69 |
| LV cardiac output (L/min) | -0.3 ± 0.9 | -0.3 ± 1.5 | 0.85 |
| LV myocardial mass (g) | 0.3 ± 6.2 | -2.6 ± 7.8 | 0.01 |
| Global peak strain Ecc (%) | -0.1 ± 2.3 | -0.4 ± 2.7 | 0.51 |
| Global peak strain Err (%) | 0.1 ± 6.7 | -0.0 ± 5.8 | 0.89 |
| Global peak strain Ell (%) | -0.3 ± 2.4 | -1.0 ± 2.3 | 0.09 |
| **Right ventricle** |  |  |  |
| RV end-diastolic volume (mL) | -0.8 ± 14.9 | -0.9 ± 26.1 | 0.95 |
| RV end-systolic volume (mL) | 0.1 ± 8.5 | 0.7 ± 12.9 | 0.72 |
| RV stroke volume (mL) | -0.9 ± 12.9 | -1.6 ± 18.8 | 0.76 |
| RV ejection fraction (%) | -0.3 ± 4.9 | -0.6 ± 5.5 | 0.75 |
| **Left atrium** |  |  |  |
| LA maximal volume (mL) | -0.8 ± 15.6 | 1.5 ± 18.1 | 0.43 |
| LA minimal volume (mL) | 0.8 ± 11.3 | 1.5 ± 9.8 | 0.74 |
| LA stroke volume (mL) | -1.6 ± 8.8 | 0.0 ± 10.5 | 0.33 |
| LA ejection fraction (%) | -1.4 ± 9.2 | -1.0 ± 6.0 | 0.82 |
| **Right atrium** |  |  |  |
| RA maximal volume (mL) | -0.4 ± 17.8 | 2.2 ± 22.8 | 0.45 |
| RA minimal volume (mL) | 1.4 ± 11.8 | -1.4 ± 14.7 | 0.21 |
| RA stroke volume (mL) | -1.8 ± 10.5 | 3.6 ± 18.9 | 0.01 |
| RA ejection fraction (%) | -1.5 ± 8.5 | 2.1 ± 10.3 | 0.02 |
| **Ascending aorta** |  |  |  |
| AAo maximal area (mm<sup>2</sup>) | 30.5 ± 54.9 | 17.8 ± 39.0 | 0.47 |
| AAo minimal area (mm<sup>2</sup>) | 34.8 ± 51.6 | 16.0 ± 41.6 | 0.26 |
| AAo distensibility (10<sup>-3</sup> mmHg<sup>-1</sup>) | -0.3 ± 1.2 | -0.0 ± 0.5 | 0.48 |
| **Descending aorta** |  |  |  |
| DAo maximal area (mm<sup>2</sup>) | 12.5 ± 29.6 | 7.0 ± 16.7 | 0.56 |
| DAo minimal area (mm<sup>2</sup>) | 15.1 ± 28.7 | 8.5 ± 23.4 | 0.47 |
| DAo distensibility (10<sup>-3</sup> mmHg<sup>-1</sup>) | -0.3 ± 1.5 | -0.1 ± 0.7 | 0.57 |

**Table S4.** Regression models to investigate the effect of SARS-CoV-2 infection-related hospitalisation on the longitudinal changes of cardiac imaging phenotypes amongst cases. Model 1: ΔIDP ∼ Sex + ΔAge + ΔBMI + ΔSBP + COVID hospitalisation; Model 2: ΔIDP ∼ Sex + Age_2_ + ΔAge + ΔBMI + ΔSBP + Cardiac disease + Asthma + Diabetes + COVID hospitalisation. Regression is applied to COVID cases. The columns report the regression coefficients and the p-values for COVID hospitalisation. The Bonferroni threshold for multiple comparison (27 imaging phenotypes) is p_Bonf_ = 0.0019 for α = 0.05. Bold font indicates a p-value < p_Bonf_.

|  | Model 1 |  | Model 2 |  |
| --- | --- | --- | --- | --- |
|  | beta | p value | beta | p value |
| **Left ventricle** |  |  |  |  |
| LV end-diastolic volume (mL) | 0.25 | 0.93 | 0.36 | 0.90 |
| LV end-systolic volume (mL) | 1.05 | 0.57 | 1.38 | 0.46 |
| LV stroke volume (mL) | -0.79 | 0.75 | -1.01 | 0.69 |
| LV ejection fraction (%) | -0.30 | 0.77 | -0.54 | 0.61 |
| LV cardiac output (L/min) | 0.01 | 0.96 | -0.05 | 0.78 |
| LV myocardial mass (g) | -1.82 | 0.13 | -1.25 | 0.31 |
| Global peak strain Ecc (%) | -0.19 | 0.68 | -0.06 | 0.89 |
| Global peak strain Err (%) | 0.16 | 0.91 | 0.13 | 0.92 |
| Global peak strain Ell (%) | -0.67 | 0.16 | -0.49 | 0.32 |
| **Right ventricle** |  |  |  |  |
| RV end-diastolic volume (mL) | 1.15 | 0.70 | 2.45 | 0.43 |
| RV end-systolic volume (mL) | -0.26 | 0.88 | 0.26 | 0.88 |
| RV stroke volume (mL) | 1.41 | 0.58 | 2.19 | 0.40 |
| RV ejection fraction (%) | 0.66 | 0.49 | 0.73 | 0.45 |
| **Left atrium** |  |  |  |  |
| LA maximal volume (mL) | 3.49 | 0.26 | 3.75 | 0.24 |
| LA minimal volume (mL) | 1.10 | 0.62 | 1.85 | 0.42 |
| LA stroke volume (mL) | 2.39 | 0.16 | 1.90 | 0.28 |
| LA ejection fraction (%) | 0.37 | 0.83 | -0.68 | 0.70 |
| **Right atrium** |  |  |  |  |
| RA maximal volume (mL) | 4.90 | 0.16 | 6.36 | 0.08 |
| RA minimal volume (mL) | -1.58 | 0.50 | -0.57 | 0.81 |
| RA stroke volume (mL) | 6.47 | 0.002 | 6.93 | **0.0016** |
| RA ejection fraction (%) | 3.51 | 0.04 | 2.99 | 0.08 |
| **Ascending aorta** |  |  |  |  |
| AAo maximal area (mm<sup>2</sup>) | -1.90 | 0.91 | 7.20 | 0.68 |
| AAo minimal area (mm<sup>2</sup>) | -5.98 | 0.70 | 4.58 | 0.77 |
| AAo distensibility (10<sup>-3</sup> mmHg<sup>-1</sup>) | 0.10 | 0.78 | 0.09 | 0.83 |
| **Descending aorta** |  |  |  |  |
| DAo maximal area (mm<sup>2</sup>) | -3.41 | 0.73 | 3.93 | 0.69 |
| DAo minimal area (mm<sup>2</sup>) | -2.38 | 0.80 | 5.45 | 0.56 |
| DAo distensibility (10<sup>-3</sup> mmHg<sup>-1</sup>) | 0.10 | 0.83 | 0.09 | 0.85 |

